# Off-label drug database

**DOI:** 10.1101/2021.11.25.21266864

**Authors:** J. Grosjean, C. Letord, I. Zana, E. Advenier-Iakovlev, C. Duclos, MO. Krebs, J. Charlet, SJ. Darmoni

## Abstract

**Background:** Many drugs are still being prescribed in a “off-label mode” and especially in psychiatry. Off-label prescription situations may vary depending on several factors and such practice is not well identifiable in the literature. Methods: A new public academic drug database has been recently created and is able to contain off-label indications, especially in psychiatry in the context of the PSYHAMM French research project. For each situation, bibliographic references have been collected to make the scientific information available to all. Results: this new off-label drug database contains more than 18,154 lines. It is freely available at https://www.hetop.eu/hetop/medicaments. Several off-label usages have been formally described and the system is extensible to all drugs and all specialties. Conclusion: An off-label drug database can be a valuable tool for health professionals and students.

## 1 Introduction

Prescribing off-label drugs in medicine is still a reality in 2021 [1], including for the COVID-19 [2]. Prescribing off-label drugs may be more common in rare diseases [3]. Since 2018, the LIMICS lab and the Saint Anne Hospital in Paris received a grant from the French National Research Agency to study off-label drug prescription in psychiatry (PSYHAMM project - http://www.cismef.org/psyhamm). Off-label prescribing of psychiatric drugs is common, despite lacking strong scientific evidence of efficacy and potentially increasing risk for adverse events [4]. In that context, psychiatry is one the main medical specialty, which prescribes the most off-label drugs in particular in the (university) hospitals [4].

Three kinds of off-label situations are described: (a) the most common one is the drug prescription indication, which is different from the ones that have a label from a National or European Medicine Agency (EMA) (e.g. prasozine for the treatment of post-traumatic stress disorders (PTSD), or acetylcysteine for the treatment of schizophrenia, dopaminergic agonists for the treatment of treatment-resistant depressive disorder); (b) the second situation is the prescription of a drug provided to a patient, where his/her age is inappropriate (e.g. use of duloxetine in children); (c) the third situation is the prescription of a drug, where the dosage or the way of administration is off-labelled (e.g. intravitreal injection of Avastin) [5]. Therefore, new clinical studies should evaluate the interest and the risks of these off-label prescriptions, in particular in psychiatry [6, 7]. In some cases, off-label prescriptions are based on non-covered therapy needs; in some cases, an extrapolation of the on-label is possible (“common sense prescription”). In other cases, the objective is to find a therapeutic solution to difficult clinical cases, especially in case of inefficiency (or relative lack of efficiency) of previous therapies (e.g. baclofene in case of alcohol addiction, or selegiline for the treatment of refractory depressive disorder). In France, the National Agency for the Safety of Medicines and Health Products (ANSM) is proposing a temporary guideline of off-label drug usage, when there is a strong therapeutic need and where the balance therapy/risk seems positive, based on the current scientific data published in the literature (efficiency and tolerance). Nonetheless, a majority of off-label usage in psychiatry is non-based on these guidelines [4]. Therefore, there is an urgent need to build an off-label drug database. Furthermore, the National (and Provincial) Health Agencies are eager to obtain from private and public hospitals an exhaustive list of these off-label prescriptions. This task is very difficult because of the lack of any off-label drug database, which is mandatory to obtain a common export format for these off-label prescriptions.

As far as we know, there is no published off-label drug database in the literature indexed in the PubMed bibliographic database; the following query was used:

*(((“databases as topic”[MH] OR “Data Base*”[TI] OR “databank”[TI] OR “databas*”[TI]) AND (Medication[TW] OR drug*[TW] OR “pharmaceutical preparations”[MH])) OR “databases, pharmaceutical”[MH] OR drug database*[TW]) AND (“Off-Label use”[MH] OR Unlabeled Indication*[TW] OR Off Label[TW])*

On the other hand, four drug databases contain some off-label usages: Micromedex DrugDex® [8], Wolters Kluwer Lexi-Drugs [9], AHFS drug information (for cancer uses of drugs and biologics) [10] and Elsevier [11]. Off-label drug indication data are included in these drug information databases when identified as a clinically relevant or emerging treatment by their respective editorial team.

The objective of this work is to describe the various undertaken steps to build an off-label drug database, first in psychiatry and then extended to other medical specialities.

### 2 Material and methods

During the PSYHAMM project (detection of off-label prescriptions in psychiatry), the consortium agreed to rely on published data indexed in the PubMed bibliographic database. Several queries were launched to detect off-label drug usage in psychiatry as exhaustively as possible: (a) the MeSH (Medical Subject Headings) descriptor «off-label use » was used and crossed with the following MeSH descriptors about psychiatry: “psychotropic drugs” OR “mental disorders” OR “psychiatry”. Some specific off-label drug usages were provided by the Saint-Anne Hospital. Finally, other usages were also added in the database from the pharmacoviligance notices provided by the French Drugs Agency (ANSM).

Since 2007, the Rouen University Hospital Department of Biomedical Informatics (RUH DBI) has developed a crosslingual health termino-ontology server (HeTOP; URL: https://www.hetop.eu/) containing 85 termino-ontologies in 45 languages [12]. A lot of information about drugs were existing prior to the PSYHAMM project and were integrated in the HeTOP terminology server: some information (drugs available in France and their composition) are extracted from the French Public Drug Database (BDPM) [13], provided by the ANSM. Unfortunately, these BDPM files do not always comply with drug identification standards (e.g. pharmaceutical forms), that would allow simpler use. Drug identification has been the subject of extensive standardization work, the most recent of which defines the identification of the medicinal product and the pharmaceutical product (ISO 11615, ISO 11616, ISO 20443, ISO 11238, ISO 11239, ISO 11240) in order to make international sharing of drug information possible. Analysis of existing models makes it possible to identify the concepts found in the BDPM to identify the drug: (a) namely the active substance, (b) the excipient, (c) the dose, (d) the form, (e) the route of administration, (f) the trade name, (g) the packaging, and the combination of these concepts to define clinical, pharmaceutical, medicinal and presented products. These concepts can be described using several terminologies, such as ATC from World Health Organization (WHO), MeSH (http://www.nlm.nih.gov/mesh), SNOMED (https://www.health.belgium.be/fr/terminologie-et-systemes-de-codes-snomed-ct) or UMLS. Normative standards also exist to represent forms, routes of administration (EMA Standard Terms), units (UCUM). In addition, a French interoperability standard (NF 97-555) defines in France drug identifiers (CIS, UCD, CIP), and the links between.

The BDMP makes available the files (a) of pharmaceutical specialties associating a CIS unique identifier with a name (normally to be constructed according to the model brand name, dosage, form), (b) of composition allowing to know the composition of the pharmaceutical specialty in quantity of active substance and therapeutic fraction, (c) presentations associating one or more CIP identifier(s) with one or more presentations themselves related to a pharmaceutical specialty. This information can be combined with a summary of product characteristics (SPC). The SPC is an annex to the authorization decision summarizing information, in particular on therapeutic indications, contraindications, conditions of use and adverse drug reactions. For example, “PROZAC 20 mg, capsule” is the wording of a drug product with CIS code 61885224. It contains “fluoxetine” (substance) in the form of “fluoxetine hydrochloride” (a possible salt of this substance). Its sale is authorized under the presentation “PVC-Aluminium blister pack(s) of 14 capsules”. This product presentation is referenced by a 13-digit submission identifier code (FID) on the drug box.

Finally, there is an identifier (UCD code) corresponding to the smallest element common to several presentations of the same pharmaceutical product. The UCD code represents, for each dosage form, the smallest dispensing unit (tablet, vial…). The UCD code and the number of UCDs per presentation are administered by the Inter-Pharmaceutical Club^1^.

The description of the drug by the BDMP is limited, when compared to the expectations of the different drug identification models [14]. For example, the concept “PROZAC” is not present in this database, while “PROZAC 20 mg, capsule” is present. Therefore, the RUH DBI has created a new concept named “pharmaceutical roots” corresponding to the different trade names (here “PROZAC”) present in the BDPM. These additions improve the reminder when applying a semantic annotator for drug detection in any health document. This also makes it possible to group together CISs with the same root. For example, the pharmaceutical root LEPONEX allows to bring together under the same concept two different specialties (LEPONEX 25 mg, tablet and LEPONEX 100 mg, tablet), which have the same indications (Parkinson’s disease psychosis, Treatment-resistant schizophrenia) and the same off-label usages (autistic disorder, borderline personality disorder, bipolar disorder, manic disorder, post-traumatic stress disorders, impulse control disorders).

As another example, the “Abacavir 300 mg tablet” includes the “ABACAVIR MYLAN 300 mg, film-coated scored tablet” and the “ABACAVIR SANDOZ 300 mg, film-coated scored tablet”, which are equivalent specialties.

Moreover, within the BDPM files, the routes of administration seem to be standardized, unlike pharmaceutical forms which do not seem to follow a precise model, still less packaging.

At the beginning of this project, the PSYHAMM consortium has developed a drug formal model published in [14] (see Figure 1). Based on this previous study, all the drug information was reorganised according to this drug formal model. The drug formal model [12] and the drug terminology server [14] are freely available for the scientific community.

**Figure 1.**
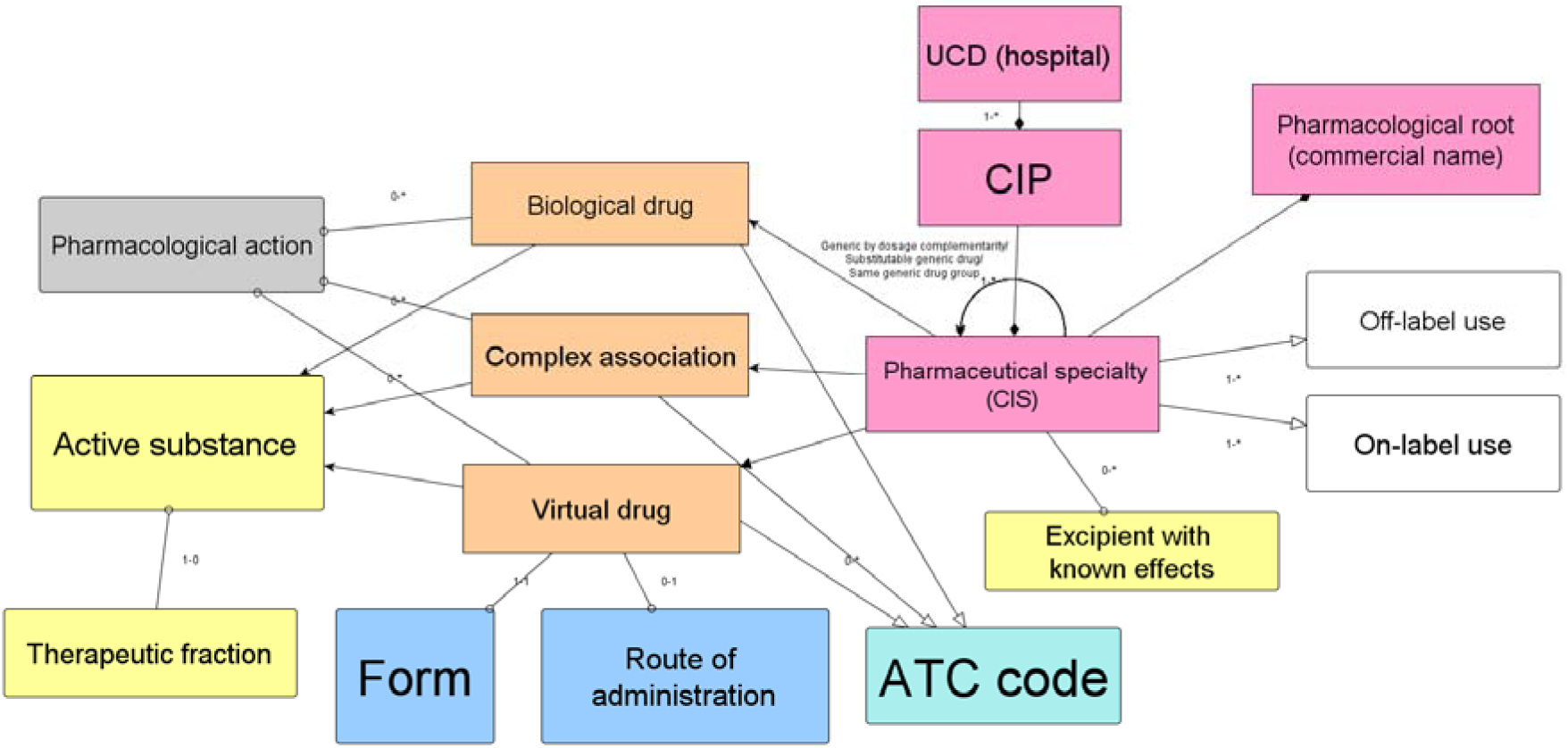
Partial drug formal model focusing on off-label

In a first step, off-label indications were linked to “drug roots”, with two potential relations: (a) in the vast majority, “therapeutic off-label indication” inspired by the MeSH qualifier “therapy”; (b) in rare cases, “off-label indication for prevention” inspired by the MeSH qualifier “prevention & control”. For these indications, as far as possible, the MeSH thesaurus was used as the reference terminology. If there was no MeSH descriptor for a specific indication, other concepts were chosen from an other terminology/ontology, with the following order of priority:

1. NCIt^2^ (National Cancer Institute Thesaurus); e.g. CATAPRESSAN (Clonidine Hydrochloride) for the treatment of “Opiate Withdrawal Syndrome”;
2. MedDRA (Medical Dictionary for Regulatory Activities); e.g. EXOMUC (acetylcysteine) for treatment and prevention of “Radiocontrast nephropathy”; and
3. SNOMED; e.g. MABTHERA (rituximab) for the treatment of “Pemphigus paraneoplastica”.

This choice was mainly based on the free access of these terminologies since MedDRA and SNOMED are under licence. If an indication did not exist in any terminology included in HeTOP, a new concept was created in a specific custom terminology.

About off-label situations linked to (a) patient age or (b) dosage or (c) route of administration, a third component has been created. This component can be complex because several situations can involve the combination of (a), (b) and (c). Thus, this data was embedded into an XML structure (using the following tags: “age-min” for the minimal age, “poso-adult” for the maximal dosage for an adult per day or week, “poso-child” for the maximal dosage for a child per day or week and “max-duration” for the maximum amount of time of the prescription (day or weeks)).

In a second step, all the off-label indications were inherited for every “drug CIS” linked via the drug information model to the “drug root”. Among these drug specialties, if one of them corresponds to an originator drug, the relationships are then inherited for each corresponding generic drug.

In a third step, in order to complete this off-label drug database, contextual bibliographic references have been added via tooltips (additional PubMed queries crossing the active ingredient and the reason for prescription) for every off-label situations. Finally, this off-label drug database is maintained in weekly basis (CL). A specific PubMed query is performed using the same MeSH Descriptor “off-label use”.

## 3 Results

Overall, this off-label drug database contains 18,154 lines. In more detail, 2,135 couples «drug roots »/indications were manually created by pharmacists (CL & AIE). Then, the following couples were automatically generated using the link drug root/pharmaceutical specialty (CIS) from the formal drug model [14]: 16,945 couples CIS/indications were generated from the previous 2,135 couples. The same pharmacists manually created: (b) 581 couples CIS/indications with maximal dosage in adults and 82 in children; (c) 465 couples CIS/indications with maximal age in children and 81 in adults. 7,170 bibliographic references are integrated in this off-label drug database (min=0; max=55 for PROLIA; median=7). Because it was integrated in the HeTOP terminology server, it follows semantic web export files, such as XML, RDF & SKOS.

The information about drugs are vast, rich and containing numerous relations, which is difficult to understand for most of the users, including clinicians. Therefore, a simpler and more compact access to drug information was created based on an HeTOP instantiation: all the drug information is accessible in a unique screen instead of three in the “regular” HeTOP. This HeTOP drug server was specifically designed for end-users, who are not experts in drug information (URL: https://www.hetop.eu/hetop/medicaments). The main use cases of this HeTOP drug server are information and teaching, whereas the “regular” HeTOP use cases are indexing (for grey literature and bibliographic database) and coding (for Diagnosis Related Groups). This HeTOP drug server as the drug information model are freely available for the scientific community or via Web services for computers.

When an end-user enters a query on HeTOP query bar, the screen will be divided in two parts. On the left, HeTOP displays the ten best candidates of the performed query. On the right, HeTOP displays all the drug information, such as different identifier codes of the drug (CIP, UCD, CIS and ATC), the components of the drug, the different pharmaceuticals specialties, pharmacological action(s), the on-label indications and even the off-label indications. As HeTOP is a terminology server and not a drug database, the end-user can click on any information to rerun a query on this new information.

Figure 2 displays an example about “PROZAC” (fluoxetine). This is the right part of the screen (presented in part 2. in the HeTOP drug description). All the different categories contain information and users can click on all the green items (metadata) to rerun a query on each chosen item. It provides all the on-label indications (n=3 in this example) and off-label indications (n=10).

**Figure 2.**
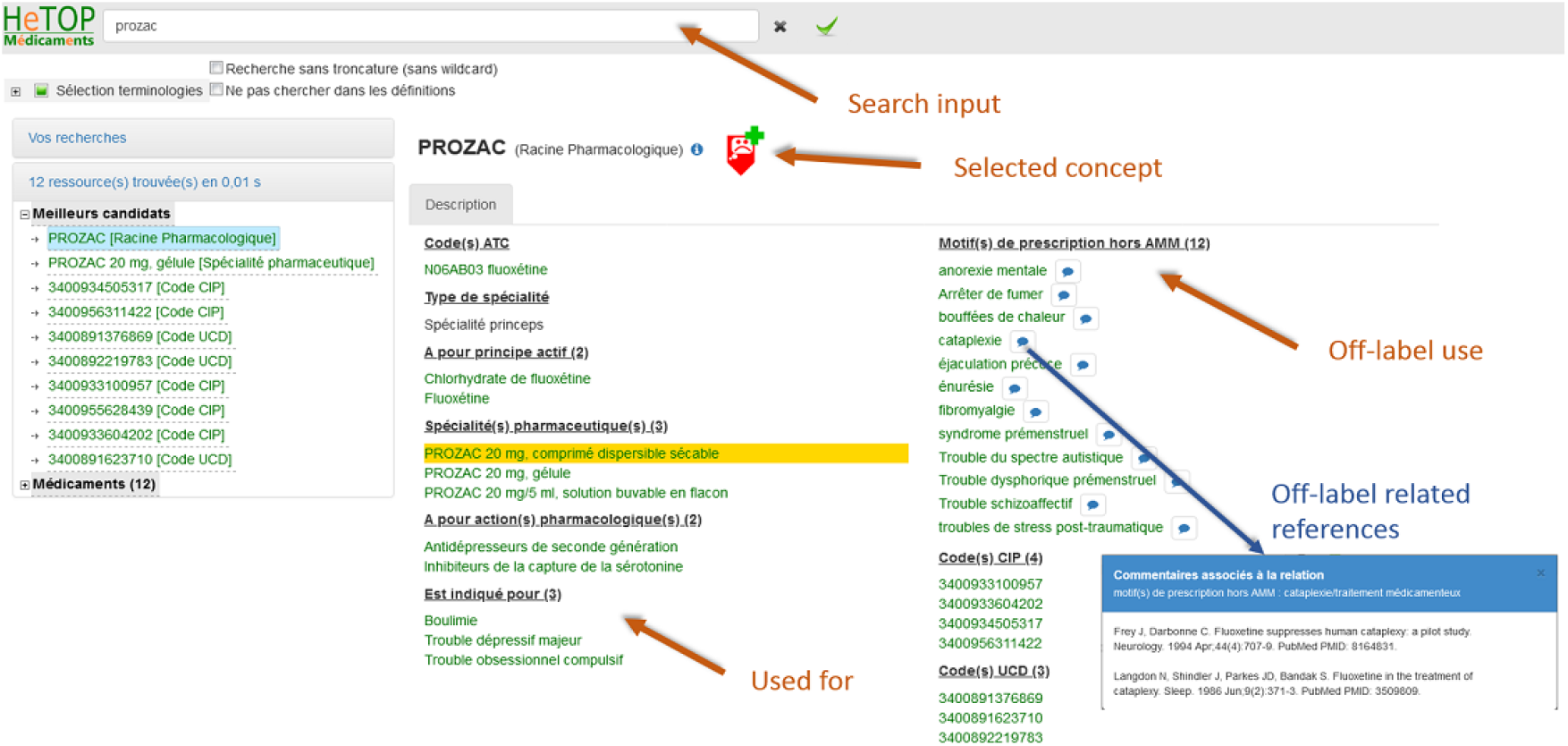
Example of drug information provided by the «HeTOP drugs » terminology server

## 4 Discussion

Although the drug is finished, identifiable, there is no universally accepted standard for naming them [5]. Depending on the point of view, it can be defined at a molecular level as an active substance, at a clinical level as a product capable of treating a pathology, at a physical level as a presentation intended to satisfy the prescription and deliverable to the patient. Drug identification can therefore be conceived at various levels with varying degrees of abstraction [6]. Other models have been adopted to represent the drug in drug databases [3] (e.g. Dictionary of Medicines and Devices from UK^3^), or to serve as a pivot between drug databases (e.g. RxNorm^4^). In these models, there is a duality between virtual and reality and the compositional relationships between active ingredients, dosage and form. These “standard” guidelines, when available, do not include French medicines. The French National

Agency for the Safety of Medicines and Health Products (ANSM) provides, via the Public Drug Database (BDPM^5^), files describing French medicines and their composition but do not always comply with drug identification standards, which would allow simple use. A Bordeaux team proposed a transformation of these files into the RxNorm format in order to have many variants naming the drugs to search for them in text reports of hospital emergency service visits [4]. In a previous work, a formal drug model was designed to link the different entities around the drug concept, but in the context of drug prescription [5].

As far as concerned, there is no academic off-label drug database published in the literature. The one created has some French specificities (in particular, some specific codes –CIP, CIS, UCD-), but the main relations between one drug and one off-label indication is available for any health professional. These relations were integrated to a drug model [14], which was extended to the entire pharmacopeia, with a weekly maintenance, using a specific query on PubMed bibliographic database. In December 2020, a French startup company (POSOS) has bought a licence to use this off-label drug database in its software suite, which is an indirect proof of its utility and its commercial value.

## Data Availability

All data produced in the present study are available upon reasonable request to the authors

## Acknowledgements

This work is partially granted by the French National Research Agency for the PSYHAMM project (https://anr.fr/Projet-ANR-18-CE19-0017).

## Footnotes

1 https://www.has-sante.fr/portail/jcms/c_671889/fr/certification-des-logiciels-d-aide-a-la-prescription-lap#U_671889/en/certification-of-software-d-aida-la-prescription-lap#U

2 https://ncithesaurus.nci.nih.gov/ncitbrowser

3 https://www.nhsbsa.nhs.uk/pharmacies-gp-practices-and-appliance-contractors/dictionary-medicines-and-devices-dmd

4 https://www.nlm.nih.gov/research/umls/rxnorm/.

5 http://base-donnees-publique.medicaments.gouv.fr

